# Proteomic profiling of aging brains identifies key proteins by which cognitively healthy centenarians defy their age by decades

**DOI:** 10.1101/2023.11.30.23299224

**Authors:** Andrea B. Ganz, Meng Zhang, Frank Koopmans, Ka Wan Li, Suzanne S.M. Miedema, Annemieke J.M. Rozemuller, Marc Hulsman, Netherlands Brain Bank, Philip Scheltens, Jeroen J.M. Hoozemans, Marcel J.T. Reinders, August B. Smit, Henne Holstege

**Affiliations:** Alzheimer Center Amsterdam, Department of Neurology, Amsterdam Neuroscience, Vrije Universiteit Amsterdam, Amsterdam UMC, Amsterdam, The Netherlands; Department of Molecular and Cellular Neurobiology, Center for Neurogenomics and Cognitive Research, Amsterdam Neuroscience, Vrije Universiteit Amsterdam, Amsterdam, The Netherlands; Department of Pathology, Amsterdam Neuroscience, Amsterdam UMC, Amsterdam, The Netherlands; Department of Clinical Genetics, Amsterdam Neuroscience, Vrije Universiteit Amsterdam, Amsterdam UMC, Amsterdam, The Netherlands; Amsterdam UMC, Vrije Universiteit Amsterdam, Department of Epidemiology and Biostatistics, Amsterdam Public Health Research Institute, Amsterdam, The Netherlands; Delft Bioinformatics Lab, Delft Technical University, Delft, The Netherlands; Netherlands Institute for Neuroscience, Meibergdreef 47, 1105 BA Amsterdam, The Netherlands

## Abstract

Some individuals reach extreme ages without any signs of cognitive decline. Here, we show that based on key proteins, cognitively healthy centenarians have a biologically younger brain. We compared the brain proteomic signatures of 58 self-reported cognitively healthy centenarians with 61 non-demented individuals and 91 AD patients. The abundance of 472 proteins strongly associated with AD Braak stages of which 64 were differentially regulated in centenarians. With increasing Braak stages, the abundance of toxic peptides of MAPT increased in AD patients, while these remained low in centenarians. Furthermore, the abundance of 174 proteins strongly changed with age, of which 108 were differentially regulated in centenarians. In fact, in brains from centenarians the abundances of essential proteins were representative of brains from individuals who were a median 18- and up to 28-years ‘younger’. The proteins involved represent diverse cellular processes, and suggest that maintained protein homeostasis is central in maintaining brain-health.

## Introduction

The incidence of aging-related diseases rises exponentially worldwide due to the increase of the average life expectancy over the last century ^1^. Among these, Alzheimer’s disease (AD) is one of the most prevalent and devastating ^2,3^. AD is characterized by the neuropathological hallmarks of amyloid beta (Aβ) plaques and neurofibrillary tangles (NFTs) ^4,5^. With increasing age, these neuropathological substrates are also found in the brains of non-demented individuals ^6–8^. Approximately 30-40% of elderly individuals aged 79 or above have significant levels of AD-related neuropathology, whereas only a small subset (15%) is clinically diagnosed with AD ^9,10^. At extreme ages, some cognitively healthy individuals accumulate levels of neuropathological substrates that can also be observed in AD patients. We previously found that a subgroup of the centenarians from the 100-plus Study cohort ^11^ was able to maintain the highest levels of cognitive performance, often for multiple years after reaching 100 years old, despite the accumulation of AD neuropathological change (ADNC), while others maintained cognitive health with relatively low levels of neuropathology ^8,12,13^. This variability suggests that those who maintain cognitive health at high age are either resilient against the effects of ADNC, or resistant against the accumulation of pathology.

To learn how cognitive health can be maintained, it is imperative to maximally comprehend the cellular and biomolecular mechanisms of prolonged resilience and resistance against neurodegeneration-associated proteinopathies. However, this is not straightforward, as many neurodegeneration-associated proteins also change with increasing age ^14,15^. In the aged and diseased brains, damaged proteins accumulate due to dysregulation of synthesis and degradation mechanisms, leading to loss-of-function and/or toxicity, which eventually contributes to the onset of neurodegenerative processes. We therefore set out to investigate whether centenarians differentially express specific proteins that support the prolongation of cognitive health. To this end, we compared brains from 58 centenarians with brains from 61 non-demented (ND) individuals and 91 AD patients, covering an age range between 50–102 years. We chose to explore the proteome from the middle temporal lobe (gyrus temporalis medialis, GTM2), as this is one of the most vulnerable regions for accumulation of ADNC during the process of AD.^16,17^ With this approach, we aimed to identify proteins that are differentially abundant in centenarians compared to expectations based on ADNC and age. We hypothesized that these proteins support the prolonged maintenance of cognitive health, and that they may point to molecular mechanisms that can slow down or even reverse the processes of aging and AD.

## Results

We performed mass spectrometry-based proteomic analysis of laser micro-dissected grey matter of the mid-temporal gyrus from 210 brains, including 58 brains from cognitively healthy centenarian, 61 brains from non-demented (ND) subjects and 91 brains from AD patients, covering a wide age range (Figure 1A). Characteristics of the 210 brains, including age, NIA Amyloid stage, Braak stage, *APOE* genotype, sex, and postmortem delay (PMD) are listed in Table S1. Quality and sample filtering resulted in 3,448 quantified proteins in 190 samples, i.e., 88 AD patients, 53 ND controls, and 49 centenarians. To investigate the molecular mechanisms underlying the resilience and resistance potential of centenarians, we set out to identify proteins that exhibit specific patterns in centenarian brains regarding neurodegeneration and aging. For this, we investigated (1) proteins whose abundance changed with increasing NIA Amyloid stage and Braak stage, which we used as a proxy for AD progression in the group of AD cases and ND individuals (Figure 1B and 1C), and (2) proteins that changed with age in ND controls. 472 proteins representing the union from an ANOVA and a linear regression test (Methods) showed the strongest changes in abundance with increasing Braak stages (Braak-proteins) (Figure S1 and Table S2). Using the same method, we identified 467 proteins with the strongest abundance-changes with increasing NIA Amyloid stage (AmyloidStage-proteins, Table S3). Of these AmyloidStage-proteins, 87.6% were also Braak-proteins (409/467), and 86.7% of the Braak-proteins were also AmyloidStage-proteins (409/472) (Figure S2); unique Braak-proteins (Table S4), and unique AmyloidStage-proteins (Table S5). Notably, the change in the abundance of the amyloid-beta precursor protein (APP) was identified both as part of the Braak-proteins and the AmyloidStage-proteins, and was driven purely by the differential abundance of the Aβ peptide (Figure 1D and S3).

**Figure 1.**
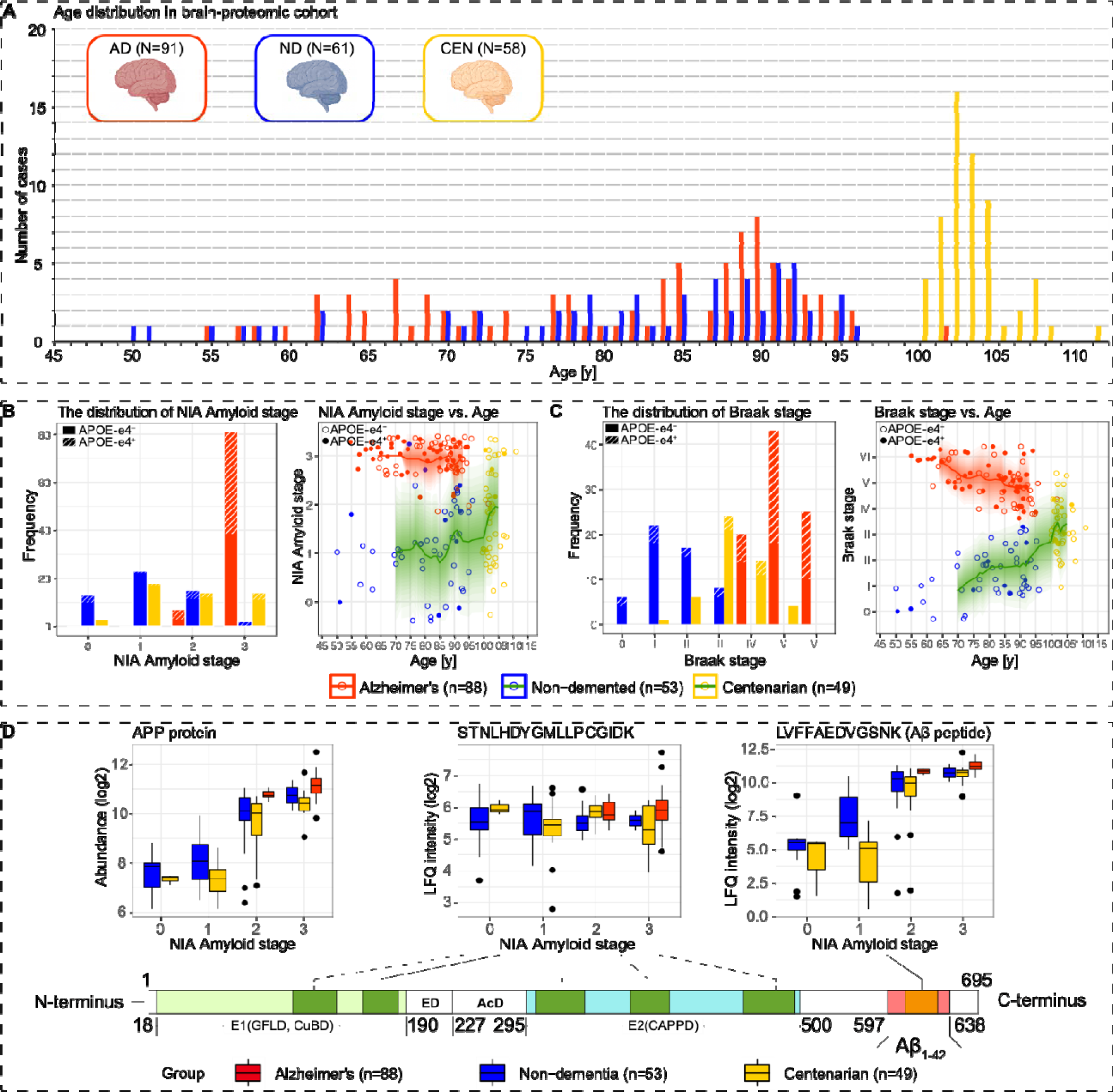
Large-scale proteomic analysis identified AD- and aging-related proteins. **A.** Age distribution of brain-proteomic samples. Red: AD cases (N=91); blue: ND controls (N=61); yellow: centenarians (N=58). **B.** The distribution of NIA Amyloid stage in filtered samples and the level of NIA Amyloid stage across age in filtered AD cases and non-demented individuals (i.e., ND controls and centenarians). Filled circles: carriers of at least one *APOE*-e4 allele. **C.** The distribution of Braak stage in filtered samples and its level across age in filtered AD cases and non-demented individuals (i.e., ND controls and centenarians). Filled circles: carriers of at least one *APOE*-e4 allele. **D.** The abundance of amyloid precursor protein (APP) and APP peptides across NIA Amyloid stages. We selected the major APP isoform in the human brain as a representative, which has a length of 695 amino acids. The extracellular region of APP is divided into the E1 and E2 domains, linked by an extension domain (ED) and an acidic domain (AcD); E1 contains two subdomains including a growth factor-like domain (GFLD) and a copper-binding domain (CuBD) interacting tightly, and E2 is the central APP domain (CAPPD). Among the measured peptides, five maintained stable abundance across the NIA Amyloid sages, including peptide STNLHDYGMLLPCGIDK; only one peptide (LVFFAEDVGSNK), which is from the Aβ1-42 fragment, increased with NIA Amyloid stages.

Braak-proteins were used as a proxy for AD. Hierarchical clustering of these proteins identified six clusters (Figure S4). EWCE cell type enrichment analysis using single-cell RNA-seq data from temporal cortical tissue as reference ^18^ and gene ontology (GO) analysis (Methods) indicated that with increasing Braak stage, we observed a decreased abundance of neuronal and mitochondrial proteins (cluster 1) and synapse-associated proteins (cluster 2). We observed an increased abundance of astrocyte- and endothelial cell-related proteins (cluster 3 and 6), with a notable enrichment for processes associated with epithelial cell differentiation, extracellular matrix (ECM), and intermediate filaments (cluster 3) and metabolic processes related to detoxification/response to oxidative stress (cluster 6). Although no specific cell types were significantly associated with cluster 4 and 5, the latter was enriched with proteins involved in processing pyridine-containing compounds, biosynthetic/metabolic processes, and glucose catabolic processes (Figure S5 and File S1).

Next, using a linear regression model on smoothed protein abundances in ND controls, we found that the abundance of 174 proteins changed significantly with age (Age-proteins), (Methods, Figure S6 and S7, and Table S6). These clustered into four unique patterns of age-related changes (hierarchical clustering, Figure S8), which were characterized using EWCE cell type enrichment analysis and GO enrichment analysis. We observed an age-associated increased abundance in cluster 1 proteins, while other clusters (cluster 2, 3, and 4) we observed a decreased abundance of proteins. Proteins in cluster 1 were enriched in excitatory neurons, in cluster 3 they were associated with oligodendrocytes and enriched for intermediate filament and myelination-related biological processes, and cluster 4 was significantly enriched in proteins associated with ribosome assembly. Notably, proteins in cluster 3 showed an accelerated decrease in abundance with age >80, indicating that oligodendrocyte-related aging may play a role after this age (Figure S9 and File S2). No specific cell types and GO terms were significantly associated with cluster 2. Notably, the abundance of many proteins in non-demented individuals have an age trajectory peaking at ∼85, which is the median age at death in the sampled population (data not shown). Because these proteins have markedly higher levels in AD patients than in non-demented individuals, and we speculate that these non-demented individuals were on a trajectory towards AD at a later age but died due to other causes.

### CEN-Braak proteins: Differential expression of 64 Braak-proteins distinguishes between brains from centenarians and AD patients with Braak stage IV

Next, we explored which Braak-proteins are differentially expressed in centenarians and AD patients, as these proteins may underlie the resilience of centenarians to increasing AD pathology. For each of the 472 Braak-proteins, we performed a two-sided t-test between the abundances observed in AD cases and centenarians with Braak stage IV. The abundance of 64 proteins (13.5% of all Braak-proteins) was significantly increased or decreased in the centenarians relative to the AD cases (CEN-Braak proteins, Table S7) with a median of 0.38 (IQR: 0.26-0.54) absolute log2-fold-change (LFC). These 64 CEN-Braak proteins are significantly enriched in cluster 2 of the Braak-proteins (28/132) (Chi-square test, p=0.002), which is enriched in neuronal cells and synapse-related biological process ontologies. This suggests that a predominant feature of cognitively healthy centenarians over AD cases with Braak stage IV is their ability to maintain synaptic functions. The 16 CEN-Braak proteins with the most significant abundance-difference across Braak stages in AD, ND, and centenarian groups are shown in Figure 2, with a complete CEN-Braak protein list in Figure S10.

**Figure 2.**
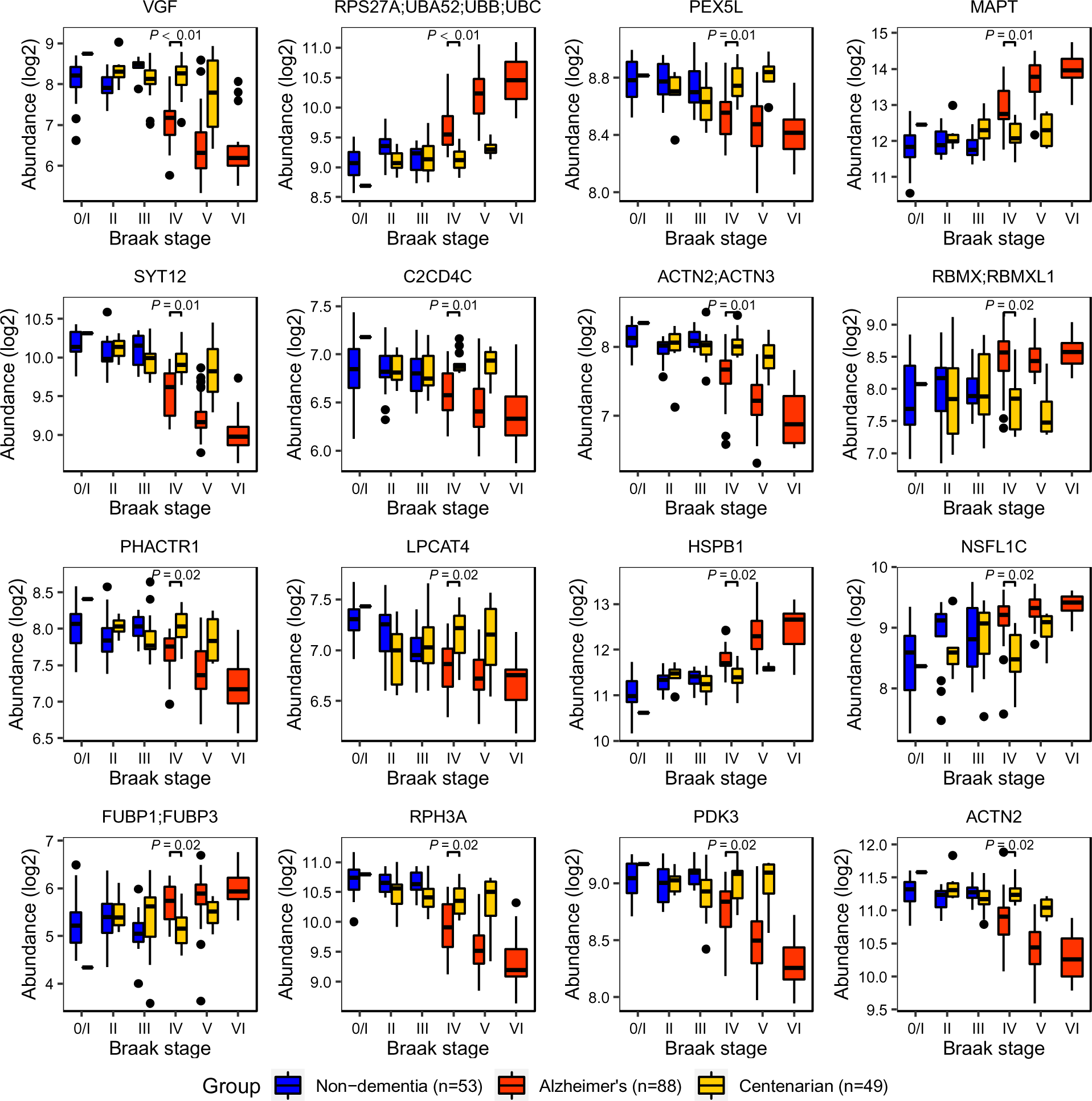
The abundance of the top-16 CEN-B proteins across Braak stages. The abundances across Braak stages in AD, ND, and centenarian groups of the top 16 proteins with the most significant p-values. The X-axis is the Braak stage from 0/I to VI; The Y-axis is the log2-transformed protein abundance.

To explore the molecular mechanisms that render the resilience to high Braak stages, we categorized the 64 CEN-Braak proteins based on their major biological functional description (Table S8). Here, we illustrate four categories: synaptic-, cytoskeleton-, microtubules-, and proteasome-related proteins (Table 1 and Figure 3); other categories are included in the supplementary file (Table S8 and Figure S11).

**Figure 3.**
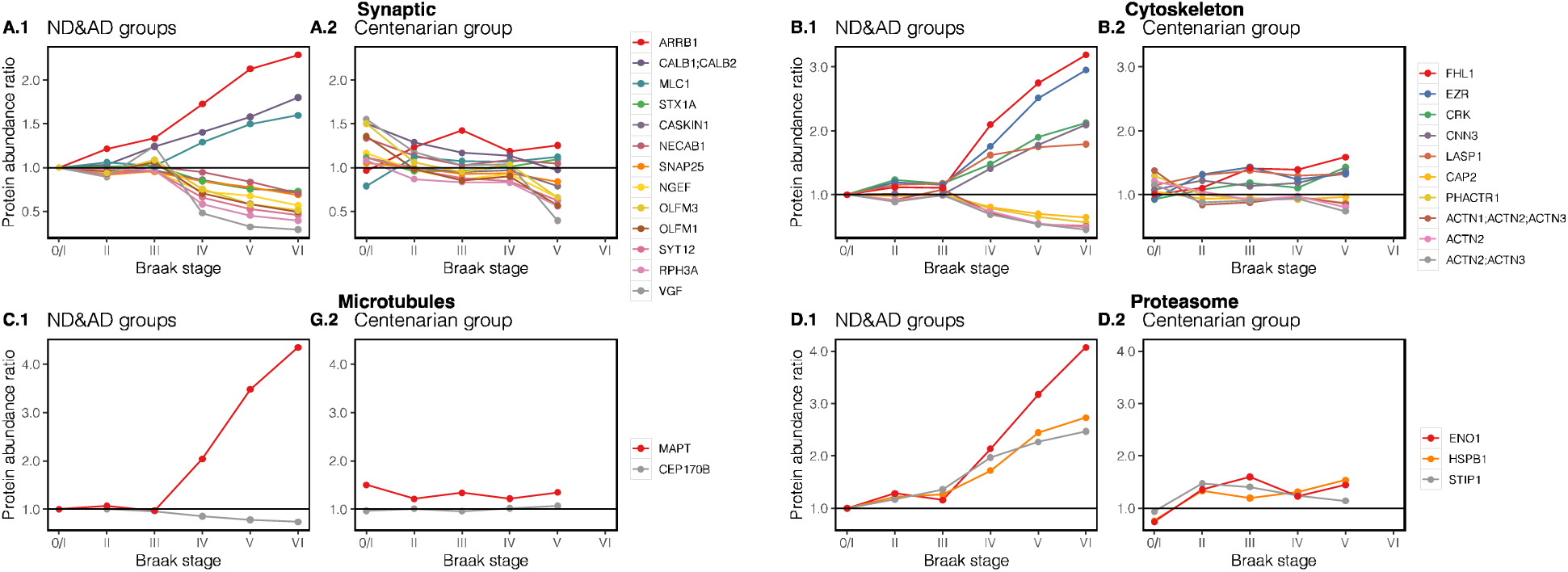
Abundance changes of CEN-B proteins vs. Braak stage. Functional protein groups are displayed along Braak stages in AD and ND groups (left panel), centenarian group (right panel). Y-axis: Protein abundance ratio: the average abundance at each Braak stage (i.e., 0/I, II, III, IV, V, VI) is depicted relative to the average abundance at Braak stage 0/I in ND individuals.

**Table 1:**
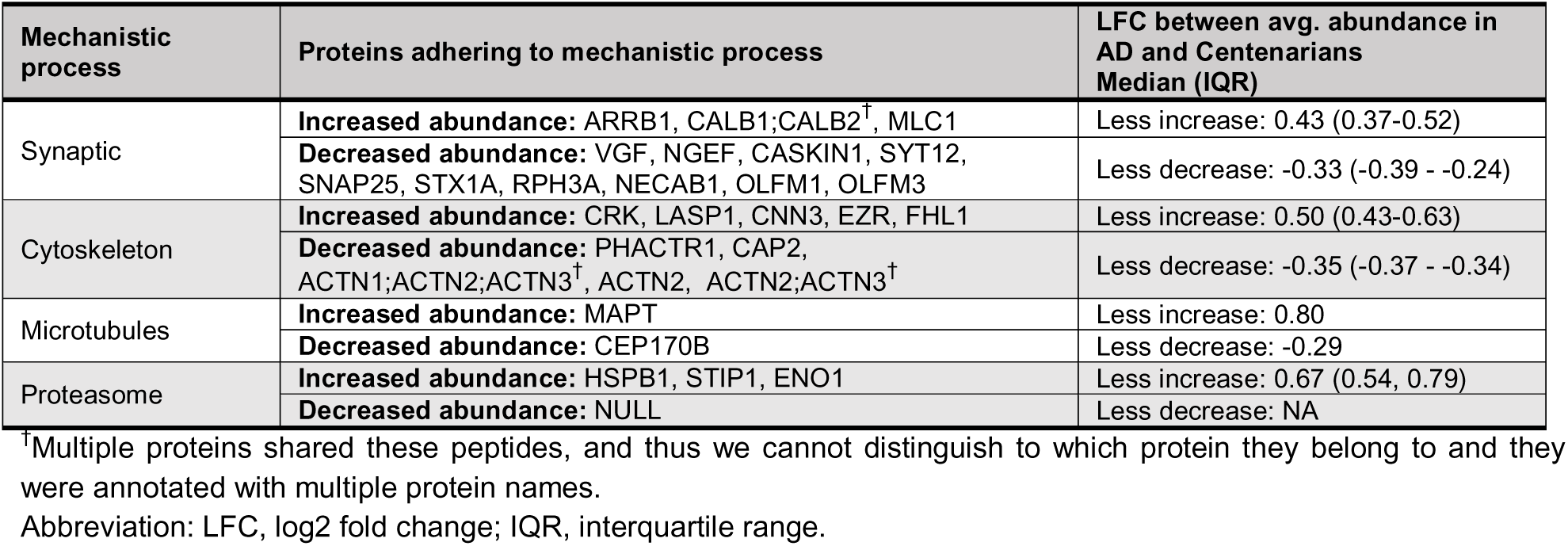
CEN-Braak protein categorizing based on their biological functions.

| Mechanistic process | Proteins adhering to mechanistic process | LFC between avg. abundance in AD and Centenarians Median (IQR) |
| --- | --- | --- |
| Synaptic | **Increased abundance:** ARRB1, CALB1;CALB2<sup>†</sup>, MLC1 | Less increase: 0.43 (0.37-0.52) |
|  | **Decreased abundance:** VGF, NGEF, CASKIN1, SYT12, SNAP25, STX1A, RPH3A, NECAB1, OLFM1, OLFM3 | Less decrease: -0.33 (-0.39 - -0.24) |
| Cytoskeleton | **Increased abundance:** CRK, LASP1, CNN3, EZR, FHL1 | Less increase: 0.50 (0.43-0.63) |
|  | **Decreased abundance:** PHACTR1, CAP2, ACTN1;ACTN2;ACTN3<sup>†</sup>, ACTN2, ACTN2;ACTN3<sup>†</sup> | Less decrease: -0.35 (-0.37 - -0.34) |
| Microtubules | **Increased abundance:** MAPT | Less increase: 0.80 |
|  | **Decreased abundance:** CEP170B | Less decrease: -0.29 |
| Proteasome | **Increased abundance:** HSPB1, STIP1, ENO1 | Less increase: 0.67 (0.54, 0.79) |
|  | **Decreased abundance:** NULL | Less decrease: NA |

<sup>†</sup>Multiple proteins shared these peptides, and thus we cannot distinguish to which protein they belong to and they were annotated with multiple protein names. Abbreviation: LFC, log2 fold change; IQR, interquartile range.

#### Synaptic proteins

Delayed changes were observed for synaptic proteins (Figure 3A): e.g., VGF Nerve Growth Factor (VGF), Olfactomedin 1 and 3 (OLFM1 and OLFM3), CASK Interacting Protein 1 (CASKIN1), and members of the SNARE complex Syntaxin-1A (STX1A), Synaptosome Associated Protein 25 (SNAP25); proteins which were previously reported to associate with cognitive performance and neuropathology in older humans ^19,20^. We also observed STX17 (Figure S11), another SNARE complex member which has been associated with autophagy ^21^. We speculate that maintaining synaptic transmission may have a protective effect against cognitive decline.

#### Cytoskeleton and microtubule-associated proteins

Centenarians maintained the abundances of cytoskeleton and microtubule-associated proteins (Figure 3B and 3C). Among these, the abundance of proteins FHL1, EZR, CRK, CNN3, LASP1, and MAPT increased with the Braak stage in AD brains but remained low in centenarians. In contrast, the levels of CAP2, PHACTR1, ACTN2, and CEP170B decreased with Braak stage in AD but maintained abundance in centenarians. Indeed, neurons are particularly susceptible to microtubule defects and deregulation of the cytoskeleton is considered to be a common insult during the pathogenesis of AD.^22^ Centenarians may be resilient to high Braak stages by maintaining the stabilization of the cytoskeleton and microtubules.

#### Proteasome

The abundance of proteasome proteins: ENO1, HSPB1, and STIP1 increased with Braak stage in AD brains but remained low in centenarian brains (Figure 3D). Low abundance of these proteins possibly reflects a non-compromised healthy proteostasis in centenarian brains.

### Different patterns were observed for MAPT peptides

The abundance of microtubule-associated protein tau (MAPT), the source protein for NFTs, deserves specific attention. In AD patients the abundance of the MAPT protein is increased for Braak stages 4, 5 and 6, however, we do not observe this increase in centenarians with the same Braak stages (Figure 4A). This difference is particularly dependent on the peptides comprising the microtubule-binding region (MTBR) in the C-terminal part of MAPT: the abundance of peptides that map to the N-terminal half of MAPT did not change with Braak stage (Figure 4B, Figure S12, and File S3). For Braak staging, NFT-*spread* across brain regions is neuropathologically assessed with Gallyas silver-staining and the AT8 antibody, to detect respectively aggregated and phosphorylated tau proteins^23^. The AT8 antibody binds the proline-rich part of MAPT at phosphorylation sites pS199, pS202 and pT205 (Figure 4B). In a subset of the cohort samples (75 AD patients, 35 ND controls, and 20 centenarians), we performed a quantitative immunohistochemistry (qIHC) analysis using the AT8 antibody, which indicated that the MAPT phosphorylation increased with Braak stage in AD cases but not in non-demented individuals and centenarians (Figure 4C). This suggests that MAPT phosphorylation is disease-associated, and may be associated with a relative increase in the abundance in the MAPT-MTBR region observed in the proteomics data. In fact, we found that the intensity of peptides from the N-terminal half showed much lower correlation with MAPT phosphorylation (Median=0.32, IQR=0.15-0.42) than peptides from the C-terminal half (Median=0.72, IQR=0.57-0.74).

**Figure 4.**
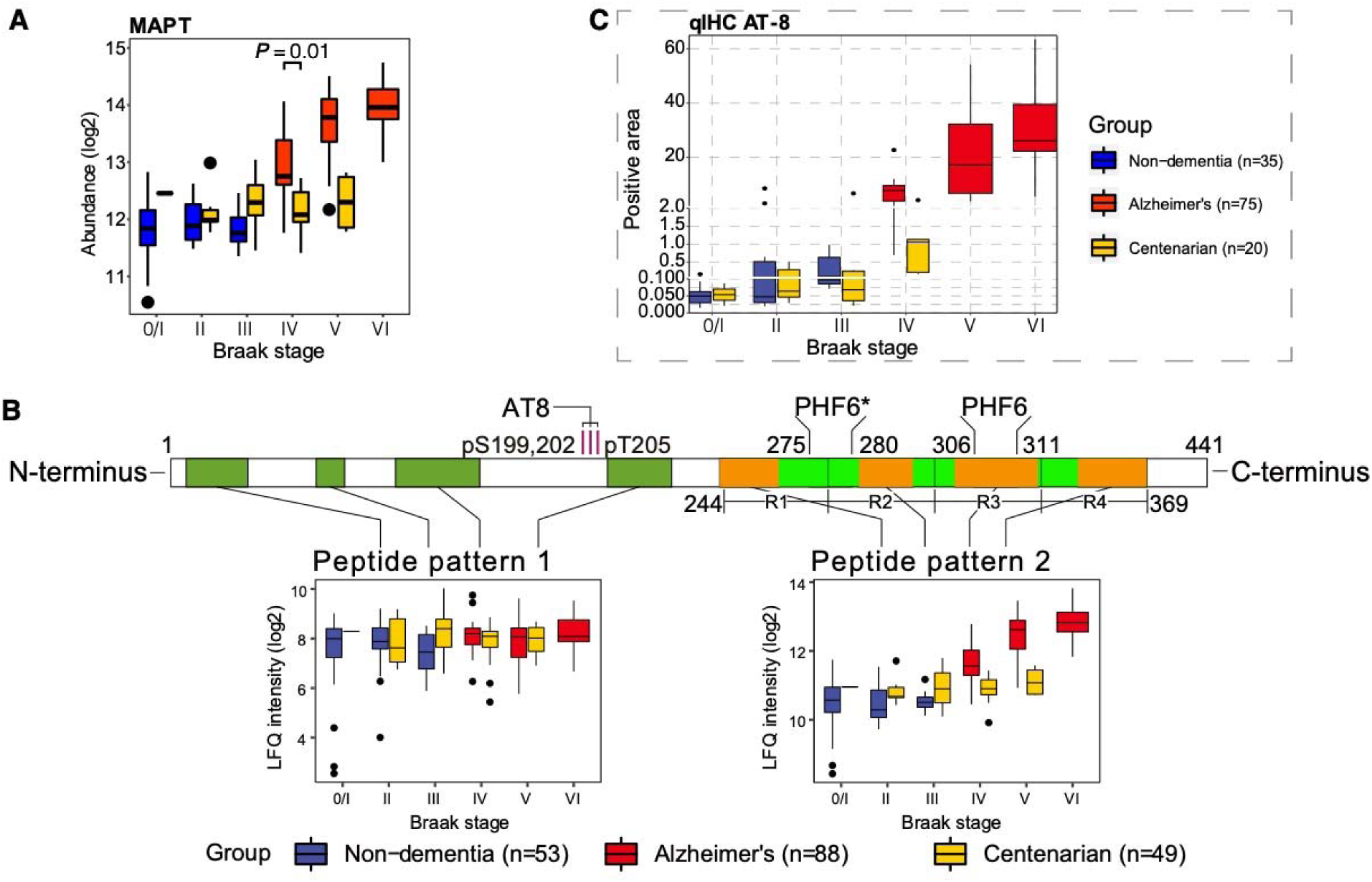
Distinguished pattern of MAPT protein in centenarian brains. **(A)** The MAPT abundances across Braak stages in AD, ND, and centenarian groups. **(B)** Peptides from the proline-rich domain of MAPT did not change with increasing Braak stages in AD nor in centenarians, whereas peptides from the microtubule-binding region (MTBR) increase with Braak stages in AD cases, but not in centenarians. **(C)** Quantified positive area of AT8 staining of a subset of the cohort (20 centenarians, 75 AD cases and 30 ND individuals) plotted against Braak stages in three different groups.

### CEN-Age proteins: Age-related proteins indicate that centenarians have a younger brain than expected for their age

The cohort design also allowed for the identification of proteins that differentiate centenarians from the ‘typical’ aging population. To do this, we compared the abundance of each Age-protein observed in centenarians to the expected abundance, by extrapolation according to their age. The expected abundance was calculated using a linear regression model, which was trained with data from ND subjects aged between 50 and 96 (Methods). Of the 174 Age-proteins, the abundance of 108 Age-proteins was significantly different between the observed abundance in centenarians and the expected abundance (CEN-Age proteins, Table S9). The protein abundance levels in centenarians were representative of protein abundances observed in individuals with much younger ages.

Using protein abundance as an estimator of biological age (Methods), we found that for these 108 proteins, the centenarian temporal cortex is estimated to be a median of 18 years (IQR:13-23) and up to 28 years younger than their chronological age. For the 66 remaining Age-proteins, for which the difference between expected and observed protein abundance in the centenarian brain was not significantly different, the protein abundance in centenarians still corresponded with abundances observed in non-demented individuals who were a median of 11 years younger (IQR:10-13) (Figure 5; Table S10).

**Figure 5.**
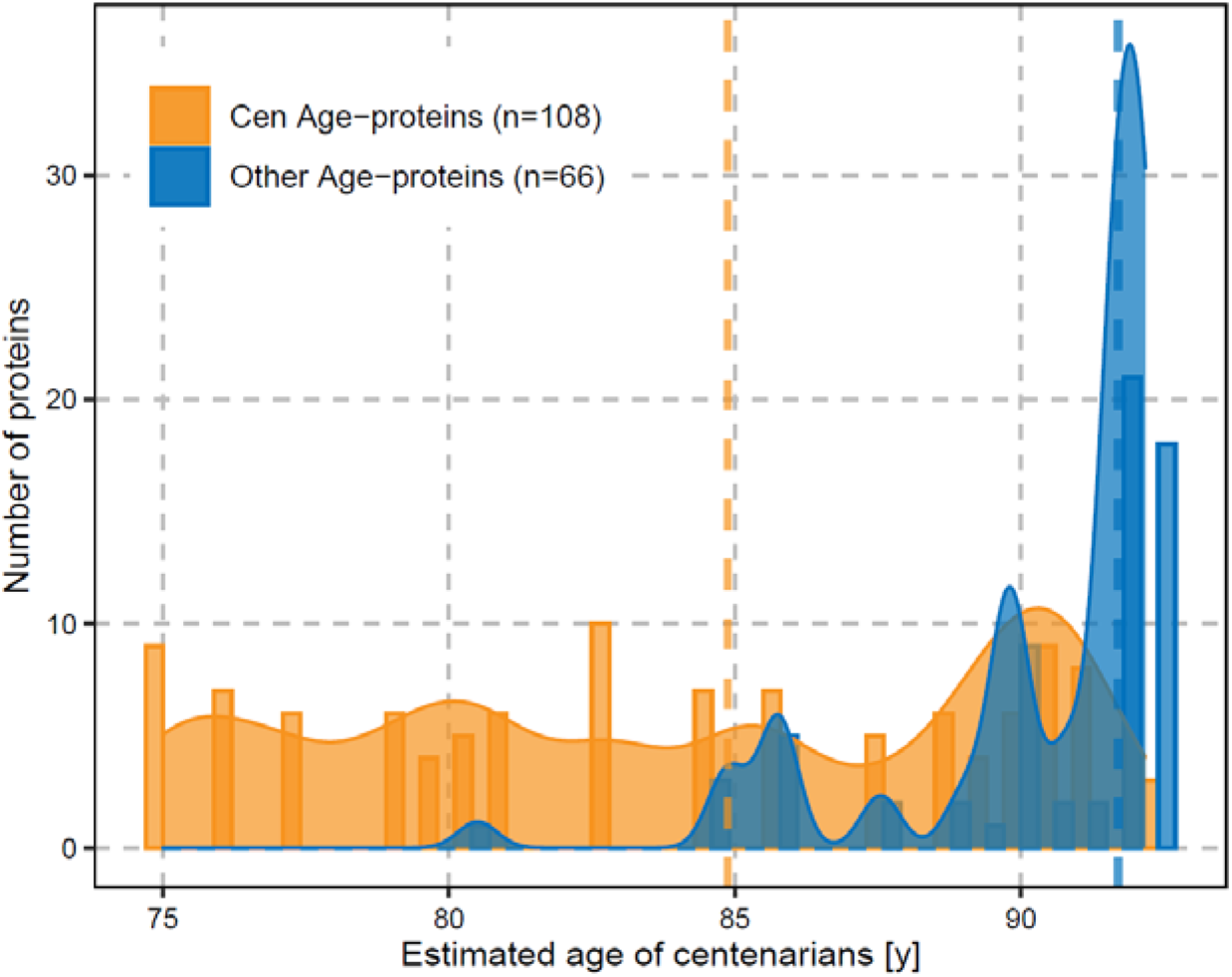
The distribution of estimated age of centenarians according to Age-proteins. Yellow bars indicate the number of CEN-Age proteins that have an abundance in centenarian brains resembling the abundance in ND brains at a certain age. The yellow curve was fitted based on the number of CEN-age proteins across ages using a Gaussian kernel. Blue bars and the blue curve are plotted the same as red bars and the red curve, but using the remaining Age-proteins. The yellow vertical line indicates the median estimated age of centenarians based on CEN-Age proteins, and the blue vertical line indicates the median estimated age of centenarians based on the remaining Age-proteins.

To explore whether these proteins might serve to uphold brain function and promote resistance to age-related decline, we assigned the functions of these 108 proteins based on manual literature curation. We identified 22 functional protein groups, and present the 9 groups associated with the strongest differences between expected and observed abundance in centenarians in Table 2. Age-dependent abundance changes (Figure 6), comprise abundance changes in proteins associated with cellular proteostasis-, cytoskeleton-, microtubules-, metabolism-, immune-response-related-, synaptic-, neurofilament-, and myelin-related proteins (the remaining 13 protein groups are in Table S8; Figure S13).

**Figure 6.**
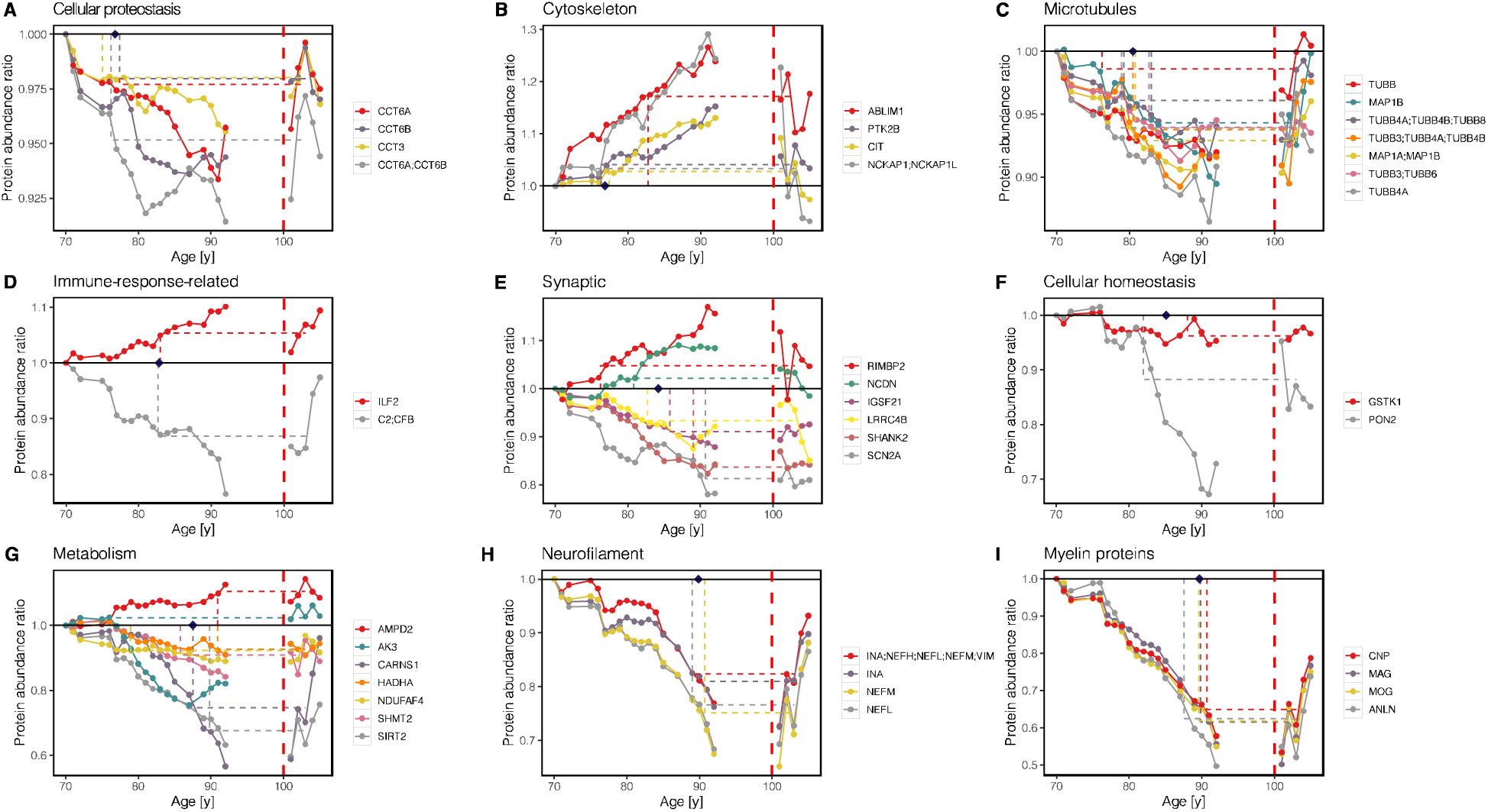
The smoothed protein abundance vs. age in ND individuals and centenarians for CEN-Age proteins. For proteins belonging to each group the smoothed protein abundances (Methods) for ND individuals and centenarians is plotted versus age. The horizontal dashed lines indicate for each protein, at which age the abundance in ND individuals is similar to the average abundance of centenarians. The blue diamond indicates the median estimated age of centenarians based on the proteins in the group. The vertical red dashed line indicates the dividing boundary between the abundance in the age continuum until 100 years, and abundance levels measured in centenarians.

**Table 2:** The abundance of CEN-Age proteins in centenarian-brains is representative of younger ages.

| Mechanistic process | Proteins adhering to a specific mechanistic process | Years younger* Median (IQR) |
| --- | --- | --- |
| Cellular proteostasis | CCT3, CCT6A;CCT6B<sup>†</sup>, CCT6A, CCT6B | 26.22 (25.66-26.54) |
| Cytoskeleton | PTK2B, CIT, ABLIM1, NCKAP1;NCKAP1L<sup>†</sup> | 26.22 (24.33-25.21) |
| Microtubules | MAP1A;MAP1B<sup>†</sup>, MAP1B, TUBB, TUBB3;TUBB4A;TUBB4B<sup>†</sup>, TUBB3;TUBB6<sup>†</sup>, TUBB4A;TUBB4B;TUBB8<sup>†</sup>, TUBB4A, | 22.54 (21.29-23.97) |
| Immune-response-related | C2;CFB<sup>†</sup>, ILF2 | 20.19 (20.11-20.26) |
| Synaptic | SCN2A, SHANK2, IGSF21, LRRC4B, RIMBP2, NCDN | 18.81 (14.85-21.76) |
| Cellular homeostasis | GSTK1, PON2 | 17.93 (15.14-20.72) |
| Metabolism | SIRT2, SHMT2, AMPD2, CARNS1, HADHA, NDUFAF4, AK3 | 15.49 (12.67-20.70) |
| Myelin proteins | MAG, MOG, ANLN, CNP | 13.34 (13.00-13.97) |
| Neurofilament | INA;NEFH;NEFL;NEFM;VIM<sup>†</sup>, INA, NEFL, NEFM | 13.19 (12.35-14.04) |

<sup>†</sup>Multiple proteins shared these peptides, and thus we cannot distinguish to which protein they belong to and they were annotated with multiple protein names.
\*The age-difference for which the average protein abundance observed in centenarians is observed in ND individuals. Abbreviation: IQR, interquartile range.

#### Cellular proteostasis

Centenarians may preserve proteostasis by maintaining the expression of proteins that belong to the T-complex protein ring complex (TRiC complex, also known as Tailless complex polypeptide 1 (TCP-1) Ring Complex). TRiC complex assists in the folding of >10% of the cytosolic proteins including actin and tubulin, and it is also obligatory for folding of many other proteins such as heat shock proteins and chaperones.^24^ While the age-related decrease in abundance of TRiC complex proteins CCT3, CCT6A, and CCT6B in ND subjects was limited, their abundances in centenarians resemble those of ND subjects who are up to 28 years younger (median: 26, IQR: 25-26), such that expression of TRIC-complex proteins are among the strongest preserved across all protein groups (Figure 6A).

#### Cytoskeleton and microtubule-associated proteins

The next best maintained proteins are cytoskeleton-related proteins (PTK2B, CIT, ABLIM1, and NCKAP1/NCKAP1L), with abundances observed in brains of individuals who were also a median of 26 years younger (Figure 6B). Centenarians resisted the age-related increase of actin binding LIM protein 1 (ABLIM1), which mediates interactions between actin filaments and cytoplasmic targets ^25^. Likewise, they maintained the abundance of protein tyrosine kinase 2 beta (PTK2B) which regulates the reorganization of the actin cytoskeleton, cell polarization, cell migration, adhesion, spreading and bone remodeling^26^. The abundance of citron Rho-interacting kinase (CIT), a serine/threonine kinase that interacts with a variety of proteins involved in actin cytoskeleton rearrangement ^27^ was in accordance with levels observed in brains from 25 years younger individuals. Furthermore, we observed that the abundance of β-tubulin family proteins (TUBB and TUBB4A) and microtubule-associated protein 1B (MAP1B) in ND subjects remained high in centenarians, and in accordance with levels observed brains from 22 years younger individuals (Figure 6C).

#### Immune-response-related proteins

Centenarians also maintained levels of proteins involved in the regulation of the innate immune response e.g., complement factor B (C2/CFB) and Interleukin 2 (ILF2) (Figure 6D).

#### Synaptic proteins

Centenarians maintained high levels of synaptic proteins, but with diverse patterns. The abundance of IGSF21, SHANK2, SCN2A, and LRRC4B decreased with age in ND subjects but remained high in centenarians, while the opposite pattern was observed for the abundances of protein RIMBP2 and NCDN (Figure 6E). Immunoglobulin superfamily member 21 (IGSF21) is a neurexin2α-interacting membrane protein that selectively induces inhibitory presynaptic differentiation ^28^. SHANK2, along with SHANK1 and SHANK3 constitute a family of proteins that function as molecular scaffolds in the postsynaptic density (PSD). SHANK directly interacts with GKAP and Homer, thus potentially bridging the N-methyl-D-aspartate receptor-PSD-95-GKAP complex and the mGluR-Homer complex in synapses ^29^. Leucine Rich Repeat Containing 4B (LRRC4B) is one of the synaptic adhesion proteins, which regulates the formation of excitatory synapses ^30^. Neurochondrin (NCDN) is a cytoplasmatic neural protein of importance for neural growth, glutamate receptor (mGluR) signaling, and synaptic plasticity ^31^.

#### Cellular homeostasis and metabolism proteins

We also observed that centenarians maintained high levels of proteins associated with cellular homeostasis (PON2 and GSTK1; Figure 6F) and cellular metabolism (SIRT2, SHMT2, AMPD2, CARNS1, HADHA, NDUFAF4, and AK3; Figure 6G). Paraoxonase 2 protein (PON2) is known for its hydrolytic and antioxidant activity, which protects brain cells from toxic substances, oxidative damage and inflammation ^32^. Similarly, Glutathione S-Transferase Kappa 1 (GSTK1) is responsible for detoxifying a wide range of harmful compounds, for and breaking down reactive oxygen and nitrogen species (ROS) and reactive nitrogen species (RNS) ^33^. Carnosine synthetase (CARNS1) catalyzes the formation of carnosine, which has been experimentally shown to reduce the effects of aging by antioxidant and anti-inflammatory properties ^34,35^. Likewise, centenarians strongly resisted age-related decrease in abundance of the Sirtuin2 protein (SIRT2), an NAD+ dependent deacetylase, which regulates microtubule acetylation and controls myelination, among its substrates ^36–38^. SIRT2 is particularly interesting, because SIRT1 (which was not among the detected proteins, likely because abundance was too low) is well known for its protective effect against aging ^39^. Adenylate Kinase 3 (AK3) involved in the reversible transfer of ATP to AMP, plays a key role in energy metabolism in the brain and is thought to regulate neuronal excitability ^40^.

#### Myelin proteins and neurofilament

Furthermore, centenarians resisted the age-related decrease of proteins that play key roles in oligodendrocytes, that build and maintain the myelin sheaths of axons, and facilitate neuronal processing speed (Figure 6H). 2′,3′-cyclic nucleotide 3′-phosphodiesterase (CNP) likely protects axons by removing toxic 2′,3′-cAMP and to replace it with axonal protectant adenosine ^41^. Myelin-associated glycoprotein (MAG) and Myelin oligodendrocyte glycoprotein (MOG) are critical for the formation and maintenance of myelin sheaths ^42–46^. In line with the resistance to age-related axonal loss, centenarians also resisted decrease in neurofilaments. The abundance of neurofilament proteins (NF), i.e., NEFL, NEFM, and alpha-internexin (INA), structural components of axons and synapses^47,48^, in centenarian brains resembled the abundance observed in non-demented individuals who were 13 years younger (Figure 6I).

## Discussion

With this work we identified, for the first time, brain proteins associated with molecular processes that potentially support the preservation of cognitive health at extreme ages. The centenarians of the 100-plus Study appear to resist important aspects of cognitive decline and overall aging ^11,49^. We observed that the abundance of key proteins typically regulated with Braak stage is unchanged in centenarians, which may contribute to their resilience against AD. Moreover, we found that centenarians were delayed or escaped key age-related changes in protein-abundance, as they had abundances of these proteins representative of individuals who were a median of 18 years, and up to 28 years younger. These proteins represent diverse cellular processes, including protein folding and aggregation, cytoskeletal rearrangement, cellular metabolism homeostasis, proteasome functioning, immune signaling, synaptic and dendritic activity. Moreover, ∼13.5% of the strongest Braak stage-related proteins play a potential role in the resilience of centenarians to higher Braak stages, representative of AD. Many of these proteins were synapse-associated proteins, suggesting that the centenarian brains maintained healthy neuronal synapses and dendrite functions despite accumulation of tau.

NFT accumulation may have limited or no adverse consequences in the centenarian brain. We speculate that this may be explained by the difference in abundance of peptides representing the C-terminal half of MAPT: these are increased with Braak stages in AD cases but not in non-demented individuals and centenarians. This C-terminal half of MAPT includes the aggregation-prone MTBR region with the R1-R4 repeats and paired helical filaments PHF6 and PFH6*, that form the aggregated core of the NFT that remains upon neuronal cell death ^50^. In addition, the phosphorylation at the AT8 sites correlated with the increased abundance of the C-terminal peptides, which supports the idea that phosphorylation of these sites promotes the aggregation process of tau ^51^. Future research will have to evaluate whether the increase in Braak stages in AD cases represents a relative increase in the toxic aggregated fraction of MAPT in AD cases compared to centenarians, who may have maintained the ability to efficiently degrade these toxic forms of MAPT.

Centenarians kept a younger temporal cortex based on a maintained abundance of 62% of the age-related proteins. Interestingly, we see that CEN-specific expression differences in proteins tend to amplify even further with increasing age. The largest difference was observed for TRiC complex proteins (with levels observed in 26 years younger brains). TRiC complex proteins are part of the cellular protein folding machinery, and assist in the folding of up to 10% of the cellular proteome in the cytosol. It is obligatory for the conformational folding of many proteins including actin and tubulin, heat shock proteins and chaperones. It is also obligatory for the unfolding and translocation of unfolded or damaged proteins for proteolysis, thereby preventing protein aggregation. Interestingly, we also observed abundances in centenarians representative of younger ages for many substrates of the TRiC complex and other chaperones, chaperonins and co-chaperones ^52,53^. Decreased levels of CCT proteins may lead to the loss of the TRiC complex, a loss of proteostasis and thus aggregation of its many client proteins, including tau, amyloid, huntingtin, and α-synuclein ^53–56^. These results suggest that maintaining high levels of chaperones and chaperone-associated proteins might be a fundamental requirement for maintained cellular health.

The next best age-shift in abundance is for proteins associated with the actin and microtubule cytoskeleton, cellular homeostasis, and metabolism, supporting the general functions of brain cells. As the actin cytoskeleton is involved in various aspects of cell biology, ranging from whole cell migration and phagocytosis to exocytosis, its maintenance and functioning is highly susceptible to disruption caused by aging. In fact, aging has been found to cause not only changes in the expression of actin but also disruption of the organization and dynamics of the actin cytoskeleton, also supported by the maintained abundance of PTK2B, CIT, and ABLIM1, for which altered abundance leads to the occurrence of age-related disorders.^57^ The age-dependent decrease of TUBB and TUBB4A proteins provide support for microtubule loss as an important component of aging.^58^ Centenarians appear to resist loss of microtubules (MT) as supported by their ability to maintain the abundance of axonal, myelin and synaptic proteins at levels representative for much younger individuals^59^ Moreover, centenarians maintained the abundance of proteins that are associated with homeostasis of cellular metabolism, such as PON2, GSTK1, CARNS1, and SIRT2. In particular, carnosine synthetase (CARNS1) catalyzes the formation of carnosine, which has been shown to reduce the effects of aging by its antioxidant and anti-inflammatory properties^60^.

Furthermore, centenarians also maintained high levels of proteins associated synaptic, neurofilament, and myelin proteins, which contribute to the preservation of cognitive performance at extreme ages. Our data suggests that with age, impaired oligodendrocyte function may reduce the capacity to produce and maintain healthy myelin sheaths ^61^, leading to myelin disruption, loss or impairment of conductivity of white matter axons and ultimately to cognitive deficits ^45,62,63^. Related to this, neurofilament proteins, especially NEFL, have recently gained attention as biomarkers of neurodegeneration in CSF and plasma ^64^. Changes in neurofilament abundance are linked to changes in both grey and white matter ^65^ and the increase of NfL levels in CSF or plasma is thought to reflect the decrease neurofilament in parenchymal tissue as analyzed here. Our data suggest that the age-related decrease of neurofilament needs to be taken into account when interpreting CSF-NFL levels as a typical AD biomarker.

Our unique dataset and study approach provided a wealth of proteomic signals underlying the resistance to disease- or age-dependent cognitive decline. However, in this study, we measured the bulk proteome, which allows us to observe changes in only those proteins with sufficient abundance. Since microglia are sparse in the brain relative to other cell-types we speculate that the changes of microglia-specific, pruning- and immune-related proteins, may be underrepresented. Furthermore, bulk measurement did not allow us to directly associate cell-specific changes in protein abundance. We referred to single cell expression data from other datasets to assign cell types, but we acknowledge that scRNA expression is a limited proxy for protein expression^66^. We also highlight that protein signals may be carried by specific peptides within each protein, which may hold information of age-related protein processing, and our observations for Aβ peptides in the APP protein and the C-terminal fragment of MAPT are a proof of principle for this phenomenon. Further, while we detected strong abundance-changes of highly relevant proteins with Braak-, Amyloid- or age (e.g. C4, CD44, GRIA3 and many others), these were not discussed in this manuscript, as we focused only the subset that was differentially expressed in centenarians. Therefore, to interrogate the trajectory of each measured protein relative to Braak stage, Amyloid stage or age, the data can be referenced with the link provided in the **Online resources**. Together, we acknowledge that to release the full potential of the different molecular constellation of the centenarian brains, a proteomic analysis with increased detection depth, analyses of posttranslational modifications, correlations with other neuropathologies, and comparison with other omics datasets is warranted.^67^

## Conclusion

The centenarian brain appears biologically younger, by preserving the abundance of proteins critically associated with neurodegenerative disease. Maintained abundance of key cytoskeletal, synaptic, and myelination proteins may be crucial for the preservation of neuronal projections and synaptic connections. Additionally, centenarians have preserved the abundance of chaperones and chaperonins, which aid in proper folding and unfolding of client proteins, preventing their aggregation. Interestingly, analysis at the peptide level of APP and MAPT suggests that a lower propensity for protein aggregation in centenarians may contribute to their resistance to neurodegenerative processes. Together, with our approach to investigate the brain proteome of cognitively healthy individuals in context of an age-continuum of AD patients and healthy individuals led to the identification of key molecular mechanisms that uphold brain health until extreme ages.

## Data Availability

All data produced in the present study are available upon reasonable request to the authors.

## Acknowledgements

We thank and acknowledge all participating centenarians and their family members and the team who recruited and visited the centenarians and their family members over the years, who collected the neuropsychological data and informed them about the option for brain donation: Chandeny Bennewitz, Debbie Horsten, Elizabeth Wemmenhoven, Esther Meijer, Ilse Admiraal, Karlijn Pieterse, Kimberley van Vliet, Kimja Schouten, Linda Lorenz, Linette Thiessen, Marieke Graat, Nina Baker, Sanne Hofman, Sterre Rechtuijt, and Tjitske Dijkstra. Finally, this work would not be possible without the great collaboration of the staff of the Netherlands Brain Bank.

## Consent Statement

The study protocol was approved by the Medical Ethics Committee of the Amsterdam UMC. Informed consent was obtained from all participants. Brain donors consented to brain donation.

## Author Contribution Statement

ABG, FK, KWL, AJMR and NBB collected and performed the proteomic measurements of the brain tissues donated to the 100-plus Study. MZ, ABG, MJTR, FK and SSMM have performed the data analysis. MZ, ABG, SSMM, JJMH, MJTR, ABS, and HH have written the manuscript. PS, JJMH, MJTR, ABS and HH supervised the research. HH designed the study. MZ, ABG, FK, KWL, SSMM, ABS and HH have verified the underlying data. All authors read and approved the manuscript.

## Data Sharing Statement

The datasets generated during and/or analyzed during the current study are available from the corresponding author on reasonable request.

## Material and Methods

### Cohorts

Tissues from Alzheimer’s disease (AD) cases and non-demented (ND) individuals were selected from the brain cohort collected by the Netherlands Brain Bank (NBB, https://www.brainbank.nl/). For each brain, we investigated clinical status prior to brain donation, to ascertain non-dementia and AD dementia. In total, we included 61 non-demented individuals spanning ages 50 to 96 and Braak stages 0 to III, and 91 AD cases with Braak stages from IV to VI, of which 48 AD cases had one copy of *APOE*-L4 allele, 43 AD cases had no *APOE*-L4 allele, spanning the ages 55 to 95 and 62 to 102, respectively. The average postmortem delay (PMD) ranges from 2.0 to 12.9 h (mean 5.7 h).

Centenarian brains were donated to the 100-plus Study, a prospective cohort of centenarians in the Netherlands ^11^. Inclusion criteria include self-reported and proxy-confirmed cognitive health and proof of age above 100 years. All participants were visited yearly at their home, where neuropsychological tests were performed. Yearly visits continued until death or until participation was no longer possible. Around 30% of 100-plus Study participants agree to post-mortem brain donation and tissue was collected in collaboration with the NBB. At the time of tissue selection for the proteomics experiment, 58 centenarians aged 100 to 111 had come to autopsy and were included. The average time between the last study visit and death is 9 months (±5 months) and postmortem delay ranges from 3.4 to 12.0 h (mean 6.5 h).

Detailed information of all 210 brains analyzed in this study, their age, Amyloid stage, Braak stage and *APOE* genotype, the sex and PMD are listed in Table S1.

### Sample preparation

Fresh frozen tissue of the middle temporal lobe (gyrus temporalis medialis, GTM2) was cut in 10 µm thick sections using a cryostat and mounted on polyethylene naphthalate-membrane slides (Leica, Herborn, DE). Sections were fixed in 100% ethanol for 1 minute and stained using 1% (wt/vol) toluidine blue in H_2_O (Fluka Analytical, Buchs, Switzerland) for 1 minute. Laser micro dissection (LMD) was performed using a Leica AS LMD system (Leica, Wetzlar, Germany) to isolate 0.5 mm^3^ of grey matter tissue and collected in 30 µl M-PER lysis buffer (Thermo Scientific, Rockford, IL, USA) in 0.5 ml Eppendorf PCR tubes and stored at -80 °C until further use.

Samples were heated to 95 °C for 5 minutes and incubated in the dark with 50 mM Iodoacetamide for 30 min at room temperature. Samples were loaded on 10% Bis/Tris-polyacrylamide gels and run into the gel for 15 min at 80 V using 1.5 M Tris/Glycine SDS running buffer pH 8.3. Gels were fixed overnight and stained with colloidal Coomassie Blue G-250, before samples were cut out and small gel pieces of about 1 mm^3^ were placed in 96-well Nunc filter plates (Thermo Scientific, Rockford, IL, USA). Destaining, trypsin digestion, and peptide extraction were performed as described previously ^68^.

Collected samples were dissolved in 100 µl Mobile phase A (2% acetonitrile/0.1% formic acid) and cleaned using the OASIS filter plate (Waters Chromatography Europe BV, Etten-Leur, The Netherlands) according to the manufacturer’s instruction. We used a subset of all samples to generate a peptide library comprising 5 groups: (1) a pool of 4 young AD cases, (2) a pool of 4 young ND individuals, (3) a pool of 4 old AD cases, (4) a pool 4 old ND individuals, (5) a pool of 8 centenarians. We further fractionated the sample pools using the Pierce high-pH reversed-phase fractionation spin columns (Thermo Scientific) according to manufacturer’s instruction but using 0.1% acetic acid instead of 0.1% trifluoroacetic acid. The collected peptides were dried and stored at -20 °C until mass spectrometry analysis.

### Mass Spectrometry

#### Library generation

For the spectral library generation, we performed a data dependent acquisition (DDA) experiment using the five pooled samples (Sample preparation). Peptides were analyzed by micro LC MS/MS using an Ultimate 3000 LC system (Dionex, Thermo Scientific) coupled to the TripleTOF 5600 mass spectrometer (Sciex). Peptides were trapped on a 5 mm Pepmap 100 C18 column (300 μm i.d., 5 μm particle size, Dionex), and fractionated on a 200 mm Alltima C18 column (100 μm i.d., 3 μm particle size). The acetonitrile concentration in the mobile phase was increased from 5 to 18% in 88 min, to 25% at 98 min, 40% at 108 min and to 90% in 2 min, at a flow rate of 5 μl/min. The eluted peptides were electro-sprayed into the TripleTOF MS, with a micro-spray needle voltage of 5500 V. The mass spectrometer was operated in a data-dependent mode with a single MS full scan (350-1250 m/z, 150 msec) followed by a top 25 MS/MS (200–1800 m/z, 150 msec) at high sensitivity mode in UNIT resolution, precursor ion >150 counts/s, charge state from +2 to +5, with an exclusion time of 16 sec once the peptide was fragmented. Ions were fragmented in the collision cell using rolling collision energy, and a spread energy of 5 eV. The mass spectra were searched against the uniprot human fasta database (full proteome, release2018-04) and iRT standard peptides using MaxQuant software (version 1.6.3.4) with default settings. The MaxQuant results were imported into Spectronaut (Biognosys) and converted into a spectral library.

#### Sample analysis

Next, we measured the proteome of all 210 individuals, using data independent acquisition (DIA). The same LC gradient used by DDA was employed for DIA. The DIA protocol consisted of a parent ion scan of 150 ms followed by a selection window of 8 m/z with scan time of 80 ms, and stepped through the mass range between 450 and 770 m/z. The collision energy for each window was determined based on the appropriate collision energy for a 2^+^ ion, centered upon the window with a spread of 15 eV. The data were analyzed using Spectronaut with the default settings. Each group of eluting peptide fragments in the raw data was matched to the spectral library by Spectronaut and yielded a compound identification score for the assigned peptide. The false discovery rate (FDR) of this quality metric was provided in Spectronaut output as q-value. In total, 28,191 peptides from 4,829 unique proteins were measured in 210 proteomic profiles.

### Quality control

#### Sample filtering

The aim of sample filtering was to remove low-quality profiles. We selected high quality samples for analyses by removing samples for which the fraction of low-quality peptides (q-value ≥0.01) exceeded 34%, i.e., when the fraction of low-quality peptides increased sharply (n=19, Figure S14A and S14B). Second, we removed samples for which the distribution of peptide abundance deviated from the overall peptide abundance distribution (Kolmogorov–Smirnov distance >0.04) (n=1) (Figure S14C and S14D). After filtering, 190 proteome profiles were left for analyses.

#### Peptide filtering

To ensure the reliability of the measured protein abundances, we performed peptide filtering based on peptide quality measures. For each protein, if ≥90% of the measured samples included at least one high-quality peptide (q-value <0.01), the measurement of this protein was considered to be reliable (n=3,448 and Table 11); otherwise, it was considered unreliable (n=1,381). For protein-measurements considered reliable, the sum of the abundance of all peptides appertaining to one protein was computed, which represented the abundance of this protein across individuals. This allows for the comparison of protein abundance between individuals, but not between the abundance of different proteins within one individual. We log2-transformed protein abundance values for analyses.

#### Variance explanation analysis

We used a mixed-effect linear model from the R-package “variancePartition” to assess the percentage of protein expression variance explained by age, sex, Braak stage, NIA amyloid stage, postmortem delay (PMD), *APOE* genotype, brain weight, and data acquisition batch. PMD, sex, and *APOE* genotype explained only minute proportions of variance in the proteomic abundance profile. However, next to Braak-stage and age (the effect of which we intended to assess in our analysis), the batch explained substantial proportions of variance (Figure S15A). We removed batch effects using the “Combat” R-package. After using “Combat”, we reassessed protein expression variance using the mixed-effect linear model, which indicated that the proportion of variance explained by batches was largely removed (Figure S15B).

#### Neuropathological measurement

Neuropathological assessments of the NBB cohort and the 100-plus Study cohort were performed by the NBB, as described previously ^13^. In this study, we investigated the levels of two AD hallmarks: (1) Aβ plaques using NIA Amyloid stage^5^, and (2) NFTs using Braak stage ^23,69,70^. Brain weight was recorded during autopsy. The centenarian brains and majority of the brains in the age-continuum were evaluated by a single neuropathologist, such that interrater variability was kept to a minimum.

#### Quantitative immunohistochemistry (qIHC) of phosphorylated Tau

To assess the load of phosphorylated tau, quantitative immunohistochemistry (IHC) analysis with antibody AT8 was performed on the GTM2 tissues from a subset of postmortem brains consisting of 75 AD cases, 35 ND individuals and 20 centenarians (Table S12). The details of IHC staining and region of interest (ROI) identification are described in the Supplementary Material. Immunoreactive area (percentage) was determined using ImageJ software ^71^.

#### Correlation between protein abundance and Braak stage or NIA-Amyloid stage

As a first identification of proteins that correlate with Braak stages and NIA Amyloid stages, we calculated the Pearson correlation in (1) all non-demented individuals and all centenarians, and (2) all AD patients.

#### Braak stage-related protein analysis

Assuming that protein abundances change between Braak stages ^72^, we categorized the data from AD cases and ND individuals according Braak stages 0/I, II, III, IV, V and VI (Braak stages 0 and I were merged, because there are only a few samples with Braak stage 0). For each protein, the significance of abundance changes across the six Braak stages was determined using a one-way ANOVA test. Next, for proteins observed to be significantly changed, we identified the Braak stage with highest median protein abundance, and the Braak stage with the lowest median protein abundance, then we calculated the log2 fold changes (LFC) between these two Braak Stages. Then, we selected the intersection across the proteins with the top 20% most significant p-values and the proteins with the top 20% absolute LFC (the cutoff of 20% was set arbitrarily). Up- and down-regulation was considered separately.

Second, assuming that protein abundance either increases or decreases with Braak Stage, we performed a linear regression model, in which the six Braak stages were treated as continuous variables with equal numerical distances. Here, we selected the intersection of the proteins with the top 20% most significant p-values in the coefficient of linear regressions *and* top 20% absolute regression coefficients. Up- and down-regulation was again considered separately.

The union of the results under each assumption was termed the “*Braak stage-related proteins”*, which we will refer to as ‘Braak-proteins’ in subsequent analysis. The proteins identified under each assumption are separately presented in two volcano plots using the ggplot2 (version 3.3.5) R-package (Figure S1A and S1B). We used the VennDiagram (version 1.6.20) package in R to indicate the intersection between the proteins identified under each assumption (Figure S1C).

#### Age-related protein analysis: Age-proteins

Age-related proteins, to which we will refer as ‘Age-proteins’ were identified by applying a linear regression model to ND brain donors, i.e., correlating the level of each protein with age-at-death. To remove person-specific differences we smoothed the observed protein abundances with samples of adjacent ages. We first sorted the ND samples according to increasing age. Then, for each center datapoint of each age, we selected the five nearest datapoints on the left and right separately, and then calculated the average protein abundance from these 11 datapoints (Figure S6). The terminal ages for which there were no five samples left or right, were excluded. For each protein, we fitted a linear regression model across smoothed abundance and age. P-values of the model coefficients were corrected for multiple testing using “Bonferroni” across all proteins tested. Changes were considered significant when corrected p-values were <0.05. Proteins that change in abundance with age are presented in volcano plots using the ggplot2 package (version 3.3.5) in R (Figure S7).

#### Clustering of proteins

We used hierarchical clustering to cluster the protein abundances (Pearson correlation coefficient as distance, Ward’s method as linkage ^73^). For Braak-proteins, the clustering was performed using protein abundances observed in AD patients and ND individuals, and for Age-proteins, the clustering was performed using protein abundances observed in ND individuals. The number of clusters for proteins was defined by evaluating the height of the dendrogram.

#### Centenarian-specific Braak-related proteins: Cen-Braak proteins

For each Braak-protein, we investigated whether the protein abundances between AD cases and centenarians at Braak stage IV differed using a t-test; both groups have the same level of NFT pathology according to Braak stages, but different cognitive status. The Braak stage IV was used, as we have similar numbers of AD cases and centenarians at this stage. The p-values were corrected for multiple testing using “Benjamini & Hochberg” method, and centenarian-specific Braak-proteins, to which we will refer as ‘Cen-Braak proteins’, were assessed while adhering to a 5% FDR cut-off.

#### Centenarian-specific age-related proteins: Cen-Age proteins

To identify protein abundances that are significantly different in centenarians than would be expected based on their age, we extrapolated the abundances of each Age-protein to ages >100 years (centenarians) according to the associated regression coefficient of the fitted linear model on the non-demented individuals. Then, to identify centenarian-specific age-related proteins, ‘Cen-Age proteins’, we used a one-side t-test (FDR<0.05) to identify significant differences between observed and expected protein abundances in centenarians.

#### Protein-dependent biological age of centenarians

To estimate the protein-specific biological brain-age of the centenarians, we calculated the difference in the average protein abundances between centenarians and the younger non-demented age-continuum. For each Cen-Age protein, we grouped the non-demented individuals per 10-year age-interval. Next, we calculated the absolute difference in the average protein abundances between centenarians and non-demented individuals from each age-interval. The age-interval with the smallest absolute difference was considered to be the biological age-interval for the centenarians based on the Cen-Age protein of interest.

Next, we assigned the mean age of non-demented individuals in this age-interval as the biological age of (all) centenarians, and the difference between this age and the mean age of the centenarians represents the estimated number of years the centenarians are biologically younger than their chronological age based on this Cen-Age protein. For each Cen-Age protein, this number of years was calculated, and the median years with interquartile range (IQR) across all Cen-Age proteins indicated how many years centenarians are biologically younger than their chronological age overall based on all Cen-Age proteins.

#### Pathway enrichment analysis

Pathway analysis was performed using topGO package in Bioconductor. The classic “Fisher” test was used to calculate the p-value, and the nodeSize was set to 5. All reliably measured proteins (n=3,448) are used as background protein list. The p-values were corrected using “Benjamini & Hochberg” method, and significance of enriched gene ontology (GO) terms was assessed while adhering to a 5% FDR cutoff.

#### Cell type enrichment analysis

Cell type enrichment analysis was performed as described previously ^74^. In short, since the brain proteins were measured from the tissue of the middle temporal lobe, we used a combination of single-cell RNA-seq data of 466 cells from eight adult control donors ^75^ and single-nuclei RNA-seq data of 15,928 cells from eight adult control donors ^76^ from temporal cortical tissue to generate the normalized gene expression data and cell type label matrices, which were subsequently used for expression-weighted cell type enrichment analysis using the EWCE R-package, version 1.2.0 ^18^. In addition, if a protein that was measured from our samples showed values ≥0.5 for a certain cell type in the normalized gene expression data and cell type label matrices, it was considered a protein marker of that cell type.

